# Bias measurement in, bias results out: how an assumption free height adjusted weight model outperforms body mass index

**DOI:** 10.1101/2020.12.22.20248739

**Authors:** Megan M. Shuey, Shi Huang, Rebecca T. Levinson, Eric Farber-Eger, Katherine N. Cahill, Joshua A. Beckman, John R. Koethe, Heidi J. Silver, Kevin D. Niswender, Nancy J. Cox, Frank E. Harrell, Quinn S. Wells

**Author notes:** Corresponding author: Quinn S. Wells, Pharm.D., M.D., Departments of Medicine, Pharmacology, and Biomedical Informatics, Vanderbilt University Medical Center, 2525 West End Avenue, Suite 300, Nashville, TN, 37203, USA. Author contributions: MMS, SH, FJH, and QSW were involved in the conception, design, analysis, and interpretation of data relating to the study as well as drafting of the initial manuscript. MMS, SH, RTL, EFE, and QSW were involved in the data acquisition while KNC, JAB, JRK, HJS, NJC, and KDN were also involved in the interpretation of results. All authors were involved in the review and revision of the manuscript as well as approval for submission.

## Abstract

**Objective:** Body mass index (BMI) is the most commonly used predictor of weight-related comorbidities and outcomes. However, the presumed relationship between height and weight intrinsic to BMI may introduce bias with respect to prediction of clinical outcomes. Using Vanderbilt University Medical Center’s deidentified electronic health records and landmark methodology, we performed a series of analyses comparing the performance of models representing weight and height as separate interacting variables to models using BMI.

**Methods:** Model prediction was evaluated with respect to established weight-related cardiometabolic traits, metabolic syndrome and its components hypertension, diabetes mellitus, low high-density lipoprotein, and elevated triglycerides, as well as cardiovascular outcomes, atrial fibrillation, coronary artery disease, heart failure, and peripheral artery disease. Model performance was evaluated using likelihood ratio, R^2^, and Somers’ Dxy rank correlation. Differences in model predictions were visualized using heatmaps.

**Results:** Regardless of outcome, the maximally flexible model had a higher likelihood ratio, R^2^, and Somers’ Dxy rank correlation for event-free prediction probability compared to the BMI model. Performance differed based on the outcome and across the height and weight range.

**Conclusions:** Compared to BMI, modeling height and weight as independent, interacting variables results in less bias and improved predictive accuracy for all tested traits.

**Study Importance Questions:** *What is already known about this subject?:*

- Body mass index, derived from collected height and weight measures, is an imperfect proxy measure of body fat composition often used in medical research.

*What are the new findings in your manuscript?:*

- We demonstrate how BMI introduces complex non-uniform biases across outcome and height-weight space.

*How might your results change the direction of research or the focus of clinical practice?:*

- Modeling height and weight as separate, non-linear, interacting variables improves clinical prediction across the complete spectrum of heights and weights for all clinical out-comes.

## Introduction

Both underweight and overweight individuals are at increased risk for adverse health outcomes and mortality.^1-4^ The most common proxy-measure of body fat in the clinical setting is body mass index (BMI),^5,6^ a derived value where height and weight are assumed to act according to the fixed relation of weight divided by the square of height (kg/m^2^). While BMI has been consistently associated with multiple weight-related outcomes,^7,8^ the intrinsic assumptions of BMI may result in limitations and biases as a predictive variable.^9,10^ For example, recent work has demonstrated that BMI undervalues the predictive potential of height for blood pressure variation and the addition of height to measures such as waist circumference improves cardiometabolic risk prediction.^11-14^

Because excess weight is known to increase risk for the components of metabolic syndrome as well as cardiovascular disease,^4,15,16^ we compared BMI to a maximally flexible, interacting model of the height/weight relationship with respect to prediction of these outcomes. Specifically, we evaluated model performance for metabolic syndrome, metabolic syndrome components, and a range of cardiovascular outcomes

## Methods

### Study population

All data were extracted from a de-identified copy of the Vanderbilt University Medical Center electronic health record on 08/2019.^17^ Measures of height and weight after 18 years of age were cleaned and units harmonized based on a previous method and BMIs were calculated.^18^

Subjects were included in landmark analyses based on a prespecified three-year qualification period that required four height and weight measures separated by approximately one year (1 year ± 4 months). For each outcome, subjects were excluded if the first occurrence of the particular outcome was before or during the qualification period. Validated data extraction methods were used to define outcomes: low high-density lipoprotein (HDL < 40 mg/dL in males and < 50 mg/dL in females), elevated triglycerides (triglycerides > 150 mg/dL), hypertension, diabetes mellitus, atrial fibrillation (AF), coronary artery disease (CAD), heart failure (HF), and peripheral artery disease (PAD).^19,20^ Because waist circumference and fasting glucose are infrequently ascertained in the clinical setting, we used a modified definition of metabolic syndrome, defined as two or more of the following events: diabetes mellitus, hypertension, low HDL, or elevated triglycerides.

### Statistical analyses

Descriptive statistics were presented as count and frequencies for categorical variables and median and interquartile range for continuous variables. Comparisons were made using Pearson’s chi-squared or Wilcoxon signed-rank, as appropriate. Cox regression analyses were conducted to examine how weight and height at the end of the qualification period (i.e., t_3_) impact the hazard of developing each outcome. Analyses utilized two models: a maximally flexible model representing height and weight (each log transformed) as separate, non-linear (restricted cubic spline with 3 knots) terms and allowing for interactions; and a log transformed BMI model. All analyses were adjusted for sex, age (with restricted cubic spline with 3 knots), and race. Model performances were evaluated by likelihood ratio, R square, and Somers’ Dxy rank correlation (index of discrimination between predicted score and observed responses). We estimated event-free probability at five years across a wide range of height-weight combinations (weight range: 50-200 kg by 5kg; height 160-200 cm by 5cm [total 270 predictions]). In predicting event-free probability at five years, patient demographic characteristics were set to the population’s median age of 51.2 years of age, white race, and female sex. Heatmaps were presented to visualize predicted 5-year event-free probability and the discrepancy between the two models across a full range of heights and weights. All statistical analyses were performed with R (version 3.3.1).

## Results

The demographics of included subjects are available in Tables S1 and S2. Metabolic syndrome event frequency was 16.7% and the frequencies for individual components were 5.5% for DM, 19.5%, for hypertension, 6.3% for low HDL, and 4.9% for elevated triglycerides. For cardiovascular outcomes, the event frequencies were 9.6% for AF, 21.8% for CAD, 8.8% for HF, and 2.8% for PAD

Performances of the two body composition models for each outcome are summarized in Table 1. Briefly, the maximally flexible height*weight model had a better log likelihood ratio, R^2^, and discrimination ability (Somers’ Dxy) than BMI with a maximum difference in model performance of 47.046, 0.0008, and 0.003, respectively.

**Table 1:** Performance Statistics for Event Free Prediction Using the Maximally Flexible Log Height-adjusted Weight Interaction or Log Body Mass Index models

| Prediction Model | LR | DF | R<sup>2</sup> | DI |
| --- | --- | --- | --- | --- |
| Metabolic Syndrome |  |  |  |  |
| log height-adjusted weight interaction | 3074.44 | 15 | 0.0424 | 0.3060 |
| logBMI | 3053.02 | 10 | 0.0418 | 0.3050 |
| Diabetes Mellitus |  |  |  |  |
| log height-adjusted weight interaction | 2735.044 | 15 | 0.0431 | 0.4319 |
| logBMI | 2687.998 | 10 | 0.0423 | 0.4289 |
| High Density Lipoprotein |  |  |  |  |
| log height-adjusted weight interaction | 1102.552 | 15 | 0.0161 | 0.2645 |
| logBMI | 1084.594 | 10 | 0.0159 | 0.2636 |
| Hypertension |  |  |  |  |
| log height-adjusted weight interaction | 2900.190 | 15 | 0.0538 | 0.3221 |
| logBMI | 2877.066 | 10 | 0.0534 | 0.3214 |
| Triglycerides |  |  |  |  |
| log height-adjusted weight interaction | 1134.645 | 15 | 0.0184 | 0.2981 |
| logBMI | 1112.862 | 10 | 0.0181 | 0.2959 |
| Atrial Fibrillation |  |  |  |  |
| log height-adjusted weight interaction | 5024.154 | 15 | 0.0591 | 0.4244 |
| logBMI | 4983.270 | 10 | 0.0586 | 0.4237 |
| Coronary Artery Disease |  |  |  |  |
| log height-adjusted weight interaction | 3375.364 | 15 | 0.0481 | 0.2752 |
| logBMI | 3347.507 | 10 | 0.0477 | 0.2743 |
| Heart Failure |  |  |  |  |
| log height-adjusted weight interaction | 5486.422 | 15 | 0.0665 | 0.4390 |
| logBMI | 5470.308 | 10 | 0.0664 | 0.4380 |
| Peripheral Artery Disease |  |  |  |  |
| log height-adjusted weight interaction | 2486.560 | 15 | 0.0503 | 0.5049 |
| logBMI | 2468.629 | 10 | 0.0500 | 0.5030 |

Abbreviations: BMI, body mass index; DF, degrees of freedom; DI, discrimination index (Somers' Dxy); LR, likelihood ratio

Figures 1 and 2 (columns 1 and 2) present heat maps displaying the pattern of predicted 5-year event-free probability for each outcome across the range of height and weight measures. Similarly, heat maps were used to display discrepancies between model predictions (Predicted probability_BMI_ – Predicted probability_height*weight_) (Figures 1 and 2, column 3). Colors indicate whether the BMI model predicts higher (blue) or lower (pink) probability than the height*weight model. In addition, for each outcome we present a histogram of prediction differences (Figures 1 and 2, column 4).

**Figure 1:**
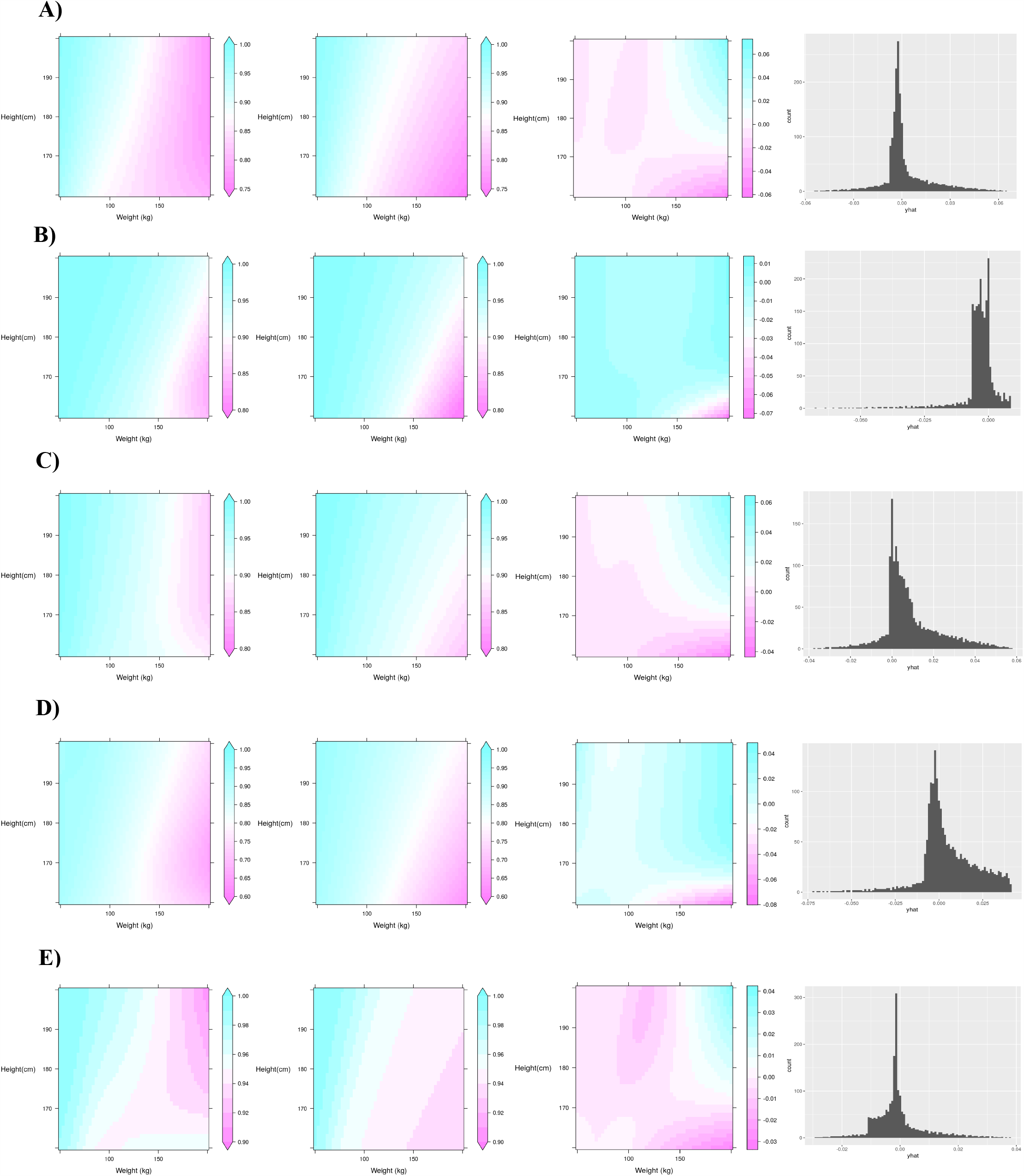
Comparison of predictive models for metabolic dysregulation and its components. A) Metabolic dysregulation; B) Diabetes mellitus; C) High density lipoprotein; D) Hypertension; E) Triglycerides. Graphics from left to right for each panel are: height-adjusted weight model prediction of 5-year free event probability across heights and weights; BMI model prediction of 5-year free event probability across heights and weights; difference in model prediction (Predicted probability_BMI_ – Predicted probability_height*weight_); distribution of the difference in the prediction difference.

**Figure 2:**
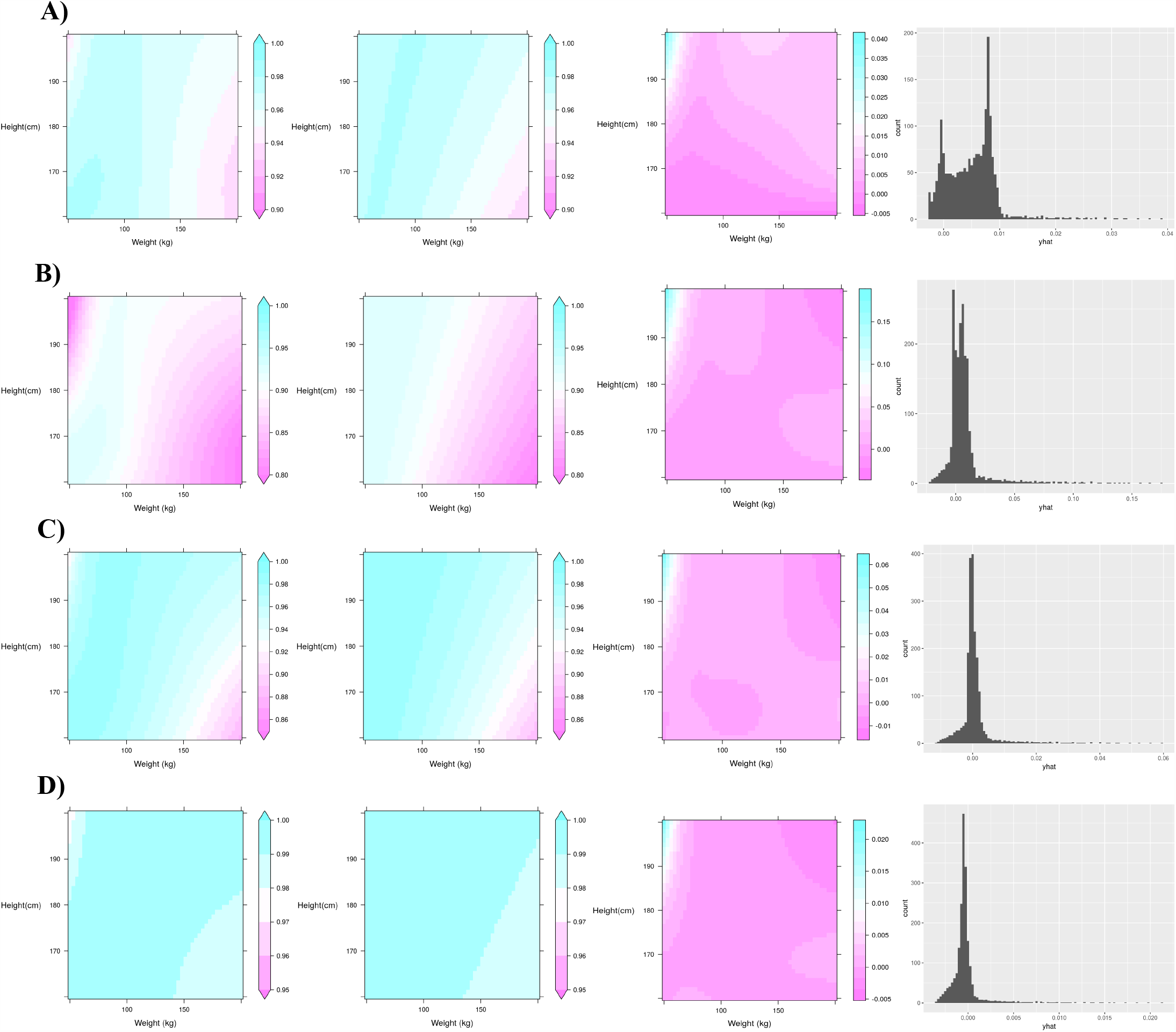
Comparison of predictive models for cardiovascular diseases. A) Atrial fibrillation; B) coronary artery disease; C) heart failure; D) peripheral artery disease. Graphics from left to right for each panel are: height-adjusted weight model prediction of 5-year free event probability across heights and weights; BMI model prediction of 5-year free event probability across heights and weights; difference in model prediction (Predicted probability_BMI_ – Predicted probability_height*weight_); distribution of the difference in the prediction difference.

## Discussion

We used electronic health record-derived data to conduct a comparative analysis of prediction models using either BMI or an unbiased height*weight interaction model. Our principal findings were that models with maximum flexibility outperform those using BMI across a wide range of cardiometabolic outcomes, and that discrepancies between models vary by outcome and location within the height-weight variable space.

Abnormal body composition is an important determinant of clinical outcomes, and accurately modeling the effect of height and weight on outcomes is critical for risk prediction. In this context, our findings have several important implications. First, although BMI is the most commonly used measure of body composition, it demonstrates inferior performance compared to a maximally flexible model across all outcomes, suggesting the assumed fixed relationship between weight and height (i.e., kg/m^2^) inadequately represents the clinical impact of body composition. Second, BMI introduces complex non-uniform biases across outcome and height-weight space. For example, predicted risk for hypertension and diabetes mellitus is similar between both approaches with the exception of high body weight individuals with short stature where BMI significantly underestimates risk. In contrast, BMI introduces considerable error into prediction of the features of atherogenic dyslipidemia (low HDL and high triglycerides), especially at higher weights, where the contribution of height is poorly modeled. Different patterns emerge for cardiovascular outcomes. BMI systematically overestimates the contribution of height for lower weight individuals and underestimates height for heavy individuals with respect to CAD, HF, and PAD risk. For instance, we observed a decrease in the predicted CAD event free risk in tall slender individuals, height > 180 cm and weight < 50 kgs, an observation that is missed by the BMI model. By comparison, BMI consistently overestimates the contribution of height for AF, though the bias is most pronounced for those at extremes of weight.

While the absolute magnitudes of discrepancies between flexible and BMI-based models were frequently modest, they are not clinically insignificant, as they are frequently not small compared to the absolute risk of the outcome in question. For example, for abnormal HDL, the BMI model overestimates the event free probability of individuals in tall patients with extreme obesity patients from 0.02 to 0.06 (2.0 to 6.0%). Considering the frequency of abnormal HDL in the total population is 6.3% this overestimate may be as frequent as the outcome alone. For abnormal HDL, a similar concern of underestimation arises in extremely heavy short patients.

The current analysis adds to the literature by systemically examining the limitations and biases of BMI across the height-weight space and a diverse set of cardiometabolic outcomes. Because BMI is calculated from height and weight, the use of BMI is a choice rather than an issue of data availability. The rationale for using BMI has often focused on the ease of calculation and interpretation, familiarity with its use among clinicians and scientists, and its established value as a predictor. Here we show that allowing height and weight to “speak for themselves”, rather than be forced to exert effects through the BMI relationship, results in more accurate risk prediction across a range of conditions and outcomes. Thus, to the extent that more accurate risk prediction translates to improved patient care, future efforts should consider more flexible approaches to modeling height and weight.

There are limitations to this study. For example, known clinical predictors for the various outcomes were not included in models. However, this choice was made to allow characterization of how choice of body weight (i.e., BMI vs height*weight) impacts model performance across outcomes. As is common with use of electronic health record data there are always concerns related to data sparsity. While our study design did its best to minimize sparsity issues it remains possible that there is potential for confounding due to this, for example it is possible that a patient may have a particular outcome, however, it was missed either due to misclassification or its development outside of the follow-up period due to various reasons.

## Conclusion

A data-driven, maximally flexible, log height-adjusted weight interaction model has better log likelihood for the prediction of weight-related outcomes than BMI. The prediction performance of these two models varies across the full spectrum of heights-weights and the absolute difference in model prediction may exceed the frequency of a given outcome. The scientific community should consider avoiding BMI when studying weight-related outcomes in favor of more flexible modeling strategies.

## Data Availability

All data is available upon request to the corresponding author.

