## Supplemental Tables for "Bias measurement in, bias results out: how an assumption free height adjusted weight model outperforms body mass index"

^*^Corresponding author

Nashville, TN, 37203, USA

**Table S1: Demographics of subjects included in time-to-event analyses for metabolic dysregulation and its components.**

|  | Diabetes Mellitus | | | High Density Lipoprotein* | | | Hypertension | | | Triglycerides^†^ | | | Metabolic Dysregulation | | |
| --- | --- | --- | --- | --- | --- | --- | --- | --- | --- | --- | --- | --- | --- | --- | --- |
|  | No  (n=84911) | Yes  (n=4945) | *P* | No (n=85021) | Yes (n=5664) | *P* | No  (n=43040) | Yes (n=10434) | *P* | No  (n=87973) | Yes  (n=4544) | *P* | No  (n=60961) | Yes (n=12187) | *P* |
| Minimum age, years | 49.7  (35.1; 62.1) | 55.3  (45.3; 64.2) | <0.001 | 51.2  (36.6; 63.1) | 50.8  (37.2; 62.0) | <0.001 | 40.2  (27.4; 53.2) | 49.9  (38.7; 59.8) | <0.001 | 51.0  (36.0; 63.2) | 51.3  (40.0; 61.3) | <0.001 | 47.2  (31.5; 60.6) | 52.3  (41.1; 62.5) | <0.001 |
| Gender, female | 53240 (62.7) | 2567 (51.9) | <0.001 | 52513 (61.8) | 3323(58.7) | <0.001 | 29777 (69.2) | 6629 (63.5) | <0.001 | 55115 (62.7) | 2486 (54.7) | <0.001 | 40970 (67.2) | 6976 (57.2) | <0.001 |
| Race |  |  | <0.001 |  |  | <0.001 |  |  | <0.001 |  |  | <0.001 |  |  | <0.001 |
| Black | 7656 (9.0) | 767 (15.5) |  | 8591 (10.1) | 670 (11.8) |  | 3135 (7.3) | 961 (9.2) |  | 20024 | 298 |  | 2189 (3.6) | 1390 (11.4) |  |
| White | 74375 (87.6) | 4082 (82.6) |  | 73618 (86.6) | 4883 (86.2) |  | 37925 (88.1) | 9193 (88.1) |  | 75108 | 4124 |  | 5320 (8.7) | 10487 (86.1) |  |
| Other | 2880 (3.4) | 96 (1.9) |  | 2812 (3.3) | 111 (2.0) |  | 1980 (4.6) | 280 (2.7) |  |  |  |  | 53452 (87.7) | 310 (2.5) |  |
| Height, cm | 167.6  (162.6;177.8) | 170.2  (162.6;177.8) | <0.001 | 167.6  (162.6;177.8) | 168.9  (162.6; 177.8) | <0.001 | 167.6  (162.6; 175.3) | 167.6  (162.6;177.0) | <0.001 | 167.6  (162.6; 177.8) | 170.2  (162.6; 177.8) | <0.001 | 167.6  (162.6;175.3) | 169.0  (162.6; 177.8) | <0.001 |
| Weight, kg |  |  |  |  |  |  |  |  |  |  |  |  |  |  |  |
| Weight t_0,_ kg | 77.7  (65.1; 92.2) | 90.0  (76.2; 105.0) | <0.001 | 78.1  (65.3; 93.0) | 86.2  (72.6; 101.7) | <0.001 | 72.6  (62.0; 87.1) | 80.0  (67.1; 94.3) | <0.001 | 78.6  (65.5; 93.9) | 85.7  (72.6; 100.2) | <0.001 | 74.8  (63.1;89.4) | 82.7  (70.3; 97.5) | <0.001 |
| Weight t_1,_ kg | 78.3  (65.5; 92.7) | 90.5  (77.1; 105.8) | <0.001 | 78.5  (65.8; 93.3) | 87.0  (72.5; 102.5) | <0.001 | 73.5  (62.6; 87.8) | 80.4  (67.6; 94.8) | <0.001 | 78.9  (65.8;94.1) | 86.4  (73.2;101.6) | <0.001 | 75.3  (63.5;89.8) | 83.5  (70.8;97.7) | <0.001 |
| Weight t_2_, kg | 78.6  (65.9; 93.2) | 90.9  (77.3; 106.6) | <0.001 | 78.9  (66.1; 93.6) | 87.5  (73.9; 103.0) | <0.001 | 74.0  (63.0; 88.5) | 81.1 (67.8;95.7) | <0.001 | 79.4  (66.2; 94.3) | 87.2  (73.8; 101.8) | <0.001 | 75.8  (64.0;90.3) | 83.9 (71.2;98.2) | <0.001 |
| Weight t_3_, kg | 78.9  (66.2; 93.4) | 91.5  (77.6; 107.0) | <0.001 | 78.9  (66.2; 93.9) | 88.5  (74.3; 103.9) | <0.001 | 74.5  (63.3; 88.9) | 81.5  (68.2; 96.2) | <0.001 | 79.4  (66.4; 94.5) | 88.0  (74.3; 102.5) | <0.001 | 76.0 (64.1; 90.7) | 84.4  (71.7;98.9) | <0.001 |
| Record length, years | 7.3  (5.1; 10.3) | 5.6  (4.1; 7.9) | <0.001 | 7.3 (5.1;10.4) | 4.6  (3.6; 6.4) | <0.001 | 6.8  (4.9; 9.8) | 5.4  (4.0; 7.8) | <0.001 | 7.27  (5.13;10.41) | 4.59 (3.59;6.34) | <0.001 | 6.7  (4.9;9.6) | 5.1  (3.3;7.3) | <0.001 |
| Days to outcome |  | 2050  (1479; 2893) | <0.001 |  | 1677  (1329; 2339) | <0.001 |  | 1979  (1442; 2831) | <0.001 |  | 167  (1310; 231) | <0.001 |  | 1851  (1393;2653) | <0.001 |

Data are presented as the median (lower; upper quartile) or number (%).

^*^ < 40 mg/dL in males and < 50 mg/dL in females

^†^ > 150 mg/dL

**Table S2: Demographics of subjects included in time-to-event analyses for cardiovascular outcomes: atrial fibrillation coronary artery disease heart failure and peripheral artery disease.**

|  | Atrial Fibrillation | | | Coronary Artery Disease | | | Heart Failure | | | Peripheral Artery Disease | | |
| --- | --- | --- | --- | --- | --- | --- | --- | --- | --- | --- | --- | --- |
|  | No  (n=86066) | Yes  (n=9094) | *P* | No  (n=54198) | Yes  (n=15074) | *P* | No  (n=86061) | Yes  (n=8321) | *P* | No (n=103239) | Yes  (n=3014) | *P* |
| Minimum age, years | 48.5  (34.5; 60.0) | 59.9  (50.1; 68.2) | < 0.001 | 44.3  (30.3; 56.9) | 53.6  (42.7; 63.6) | < 0.001 | 49.0  (34.9; 60.7) | 61.5  (51.4; 70.1) | < 0.001 | 50.7  (36.7; 62.4) | 62.7  (54.5; 70.0) | < 0.001 |
| Gender, female | 54958 (64) | 4538 (50) | < 0.001 | 36908 (68) | 9222 (61) | < 0.001 | 53676 (62) | 4353 (52) | < 0.001 | 62789 (61) | 1432 (48) | < 0.001 |
| Race |  |  | < 0.001 |  |  | < 0.001 |  |  | < 0.001 |  |  | < 0.001 |
| Black | 9272 (11) | 1029 (11) |  | 5411 (10.0) | 1804 (12) |  | 8426 (9.8) | 1039 (12.5) |  | 10644 (10) | 491 (16) |  |
| White | 73586 (85) | 7933 (97) |  | 46417 (85.6) | 12917 (85.7) |  | 74527 (86.6) | 7150 (85.9) |  | 89152 (86) | 2466 (82) |  |
| Other | 3208 (4) | 132 (2) |  | 2370 (4.4) | 353 (2.3) |  | 3108 (3.6) | 132 (1.6) |  | 3443 (4) | 57 (2) |  |
| Height, cm | 167.6  (162.6; 175.3) | 170.2  (162.6; 179.5) | < 0.001 | 167.6  (162.5; 175.3) | 167.6  (162.6; 177.8) | < 0.001 | 167.6  (162.6; 177.8) | 170.2  (162.6; 177.8) | < 0.001 | 167.6  (162.6; 177.8) | 170.2  (162.6; 178.0) | < 0.001 |
| Weight, kg |  |  |  |  |  |  |  |  |  |  |  |  |
| Weight t_0_, kg | 79.2  (65.8; 94.6) | 85.3  (71.9; 100.0) | < 0.001 | 76.7  (64.0; 92.0) | 82.1  (68.6; 97.5) | < 0.001 | 79.4  (66.1.; 94.8) | 85.3  (72.1; 100.6) | < 0.001 | 80.4  (66.9; 96.1) | 85.3  (72.4; 100.0) | < 0.001 |
| Weight t_1_, kg | 79.6  (66.2; 94.8) | 85.3  (71.9; 99.8) | < 0.001 | 77.2  (64.6; 92.5) | 82.4  (69.0; 97.5) | < 0.001 | 79.8  (66.5.; 95.0) | 85.3  (72.1; 100.7) | < 0.001 | 80.8  (67.2; 96.2) | 85.3  (72.3; 99.8) | < 0.001 |
| Weight t_2_, kg | 79.9  (66.7; 95.2) | 85.3  (72.1; 100.4) | < 0.001 | 77.7  (65.0; 93.0) | 82.5  (69.0; 98.2) | < 0.001 | 80.3  (67.0; 95.3) | 85.4  (72.2; 101.2) | < 0.001 | 81.2  (67.6; 96.6) | 85.7  (72.2; 100.2) | < 0.001 |
| Weight t_3_, kg | 80.3  (67.0; 95.5) | 85.4  (71.9; 100.3) | < 0.001 | 78.0  (65.3; 93.4) | 82.6  (69.0; 98.4) | < 0.001 | 80.4  (67.1; 95.7) | 85.7  (72.1; 101.3) | < 0.001 | 81.4  (67.9; 96.8) | 85.5  (72.1; 100.7) | < 0.001 |
| Record length, years | 7.3  (5.1; 10.44) | 6.4  (4.4; 9.2) | < 0.001 | 6.9  (4.9; 9.9) | 5.8  (4.1; 8.2) | < 0.001 | 7.2  (5.1; 10.2) | 6.0  (4.2; 8.9) | < 0.001 | 7.42  (5.2; 10.6) | 6.36  (4.5; 9.1) | < 0.001 |
| Days to outcome |  | 2321  (1608; 3346) | < 0.001 |  | 2106  (1505; 2977) | < 0.001 |  | 2185  (1544; 3259) | < 0.001 |  | 2322  (1633; 3322) | < 0.001 |

Data are presented as the median (lower; upper quartile) or number (%).
